# State-by-State estimates of R0 at the start of COVID-19 outbreaks in the USA

**DOI:** 10.1101/2020.05.17.20104653

**Authors:** Anthony R. Ives, Claudio Bozzuto

## Abstract

We estimated the initial rate of spread (*r*_0_) and basic reproduction number (R0) for States in the USA experiencing COVID-19 epidemics by analyzing death data time series using a time-varying autoregressive state-space model. The initial spread varied greatly among States, with the highest *r*_0_ = 0.31 [0.23, 0.39] (95% CI) in New York State, corresponding to R0 = 6.4 [4.3, 9.0] (95% CI). The variation in initial R0 was strongly correlated with the peak daily death count among States, showing that the initial R0 anticipates subsequent challenges in controlling epidemics. Furthermore, the variation in initial R0 implies different needs for public health measures. Finally, the States that relaxed public health measures early were not those with the lowest risks of resurgence, highlighting the need for science to guide public health policies.

---

The basic reproduction number (R0) is a fundamental metric in epidemiology that gauges how rapidly a disease will spread at the beginning of an epidemic (Delamater et al. 2019). While R0 depends in part on the biological properties of the pathogen, it also depends on properties of the host population that, for example, determine the contact rate between individuals (Hilton and Keeling 2020). Estimating R0 is a requisite for designing public health measures for infectious diseases such as COVID-19.

Computing a reliable estimate of R0 poses challenges (Obadia et al. 2012, Ma et al. 2014, Delamater et al. 2019), as illustrated by the wide range of values estimated for the COVID-19 epidemic in Wuhan, China (Table 5 in Liu et al. 2020). Although R0 can be calculated by following the chain of infections between individuals using case-tracking, calculating R0 from the population-level spread of a disease has the advantage of integrating all of the complexities of human interactions. Moreover, this potentially reveals the effects of enacted public health measures (Flaxman and al. 2020, Hurtado and Tinajero 2020).

We computed R0 at the onset of epidemics for States in the USA from data on the number of deaths per day (Fig 1A,B) (The New York Times 2020). To meet the challenge posed by changes that affect R0 and the low numbers of counts in many States, we fit the State-level time series using a hierarchical time-varying autoregressive model to estimate *r(t)* that accounts for later changes caused by public response to the epidemics (Ives and Dakos 2012, Bozzuto and Ives 2019) (Supporting Information). From *r*(*t*) we then computed R0(*t*) using the Dublin-Lotka equation (Dublin and Lotka 1925) with the probability distribution of the timing of transmission (Li et al. 2020). We focused the analysis on death data even though fatalities (fortunately) represent only a fraction of newly infected cases, because deaths are less likely to be misreported compared to data on the number of cases. The proportion of people who have recovered from COVID-19, and are consequently immune, is still likely to be small, and therefore we refer to our estimates as the time-varying R0(*t*), in contrast to the effective reproduction number, *R(t*) (Flaxman and al. 2020, Hurtado and Tinajero 2020, Systrom and Vladeck 2020).

**Fig. 1.**
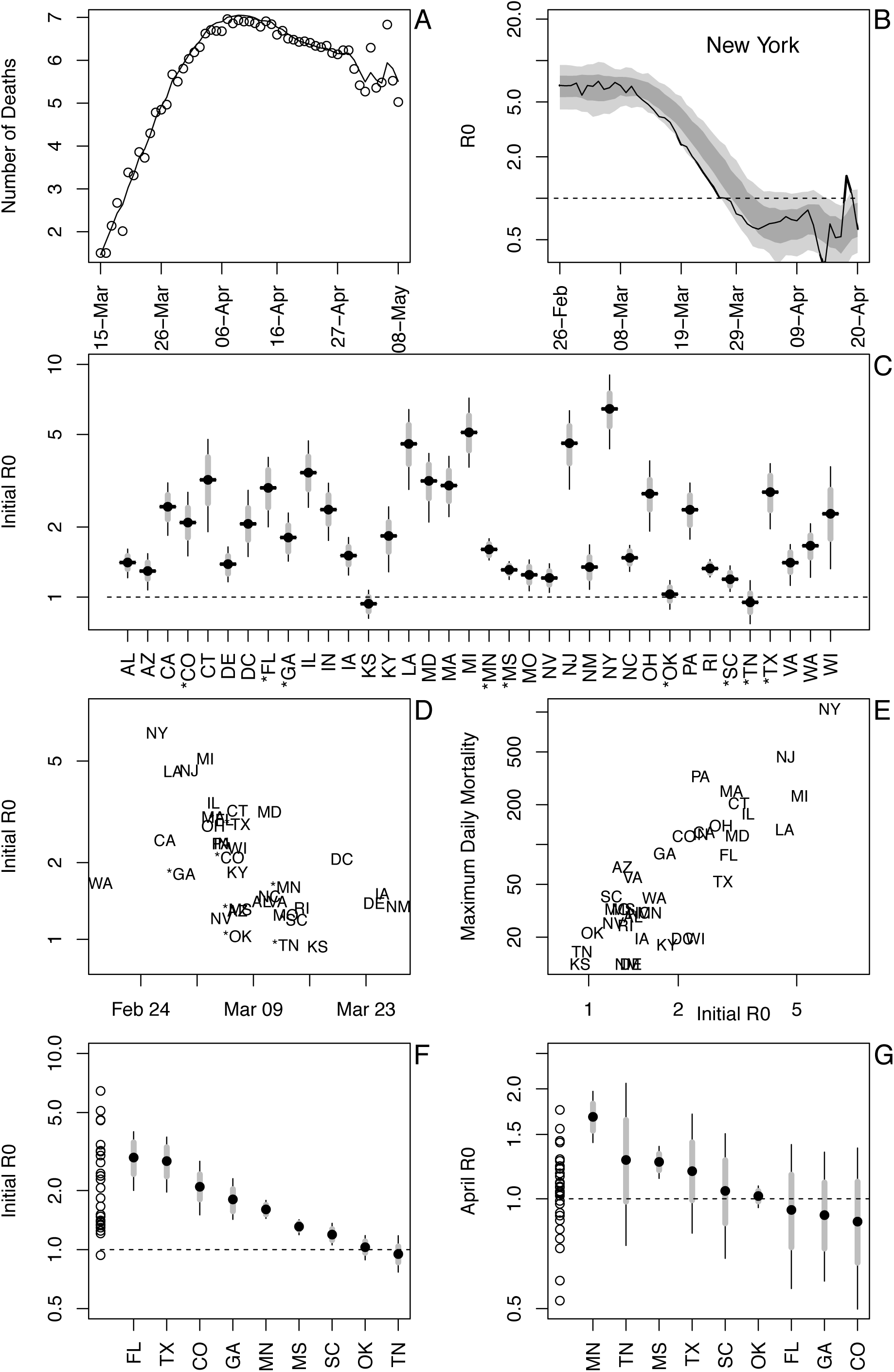
Panels **(A)-(B)** show data and model fit for New York State as an illustrative example. **(A)** Death data time series (log-transformed), along with the model fit. **(B)** Estimated time-varying R0(*t*), with point estimates (solid line) and accompanying approximate confidence intervals (66% in dark gray, 95% in light gray). The fitting uses a state-space model with time-varying spread rates and quasi-Poisson sampling of death counts, and approximate confidence intervals are obtained with parametric bootstrapping. **(C)** Estimated initial R0 values for all analyzed States, shown as bootstrapped estimates (black horizontal bars) and approximate 66% and 95% confidence intervals (boxes and whiskers). Asterisks mark nine of the eleven States that eased restrictions in April, 2020; the remaining two States did not have sufficient numbers of deaths for estimating R0. **(D)** Estimated initial R0 values ordered by the date of epidemic onset. **(E)** Maximum daily number of deaths in each State ordered by estimated initial R0 values. **(F)** Estimated initial R0 values from death data for nine of the eleven states in which restrictions were relaxed in April, 2020. For comparison, on the far left (dots) are the R0 bootstrapped estimates for all other States for which estimates were made. **(G)** Estimated R0 values using case data at the end of April, 2020, for nine of the eleven States for which we estimated R0, with R0 estimates for the remaining States on the far left.

Initial R0 values varied dramatically for those States that have experienced enough deaths for estimation (Fig. 1C). These values as a group are likely to be, if anything, underestimates due to the statistical challenges of inferring values of R0 that are changing rapidly, as they might at the start of an epidemic (Supporting Information). Estimates of the initial R0 with the same statistical method applied to reported case data show similar but generally higher values (Supporting Information).

Some of the State-to-State variation is attributable to States with later epidemic onset showing lower initial R0 values (Fig. 1D). This might be explained by measures that citizens and governments took before the epidemic started within a State: States in which the epidemic (inferred by deaths) started before the World Health Organization declared COVID-19 a pandemic (11 March, 2020) had higher estimates of R0 (t_34_ = 3.68, P = 0.0008). Nonetheless, the relationship is not strong and has clear exceptions: for example, Washington State was the first to report multiple deaths but nonetheless maintained low values of initial R0.

High values of R0 also imply high potential impacts of epidemics. This is seen among States, for which higher initial R0 values correspond to higher peak daily mortalities (Fig. 1E). This relationship is not tautological (with high mortality causing high estimates of R0), because our estimates of R0 are for the onset of epidemics.

The threat of high potential R0 values is important when assessing the risks of easing public health measures that States have enacted to slow their epidemics. In the second half of April, 2020, eleven States reduced or removed restrictions; enough deaths had occurred in nine of these States for us to estimate R0 (Fig. 1C, asterisks). As a group, they did not have lower initial R0 than other States (Fig. 1F, *t* = −1.46, *P* = 0.15). A similar analysis using case data shows that States relaxing restrictions did not have, on average, lower estimates of R0 at the end of April (Fig. 1G, *t* = 0.55, *P* = 0.58; Supporting Information). This suggests that restrictions were not removed according to either the potential risk of disease spread within States (the initial R0, Fig. 1F) or the risk of spread at the time restrictions were removed (R0 at the end of April, Fig. 1G).

The presented State-by-State estimates of R0 confirm what is widely understood: COVID-19 has the potential to spread rapidly, especially in urban areas (Fig. 1C), and States differ greatly in how they have experienced the disease. Our estimates put numbers to these observations. Estimates of R0 reveal the potential threat of epidemics when restrictions are eased, as well as the potential challenges for vaccination campaigns: using the rule of thumb that a proportion 1 − 1/R0 of a population needs to be vaccinated (but see Gerberry and Philip 2016), vaccination coverage will have to be roughly 60% on average among the States we analyzed (Fig. 1C). This, however, is misleading, because it does not consider the variation among States, and for New York State the R0 = 6.4 suggests a vaccination rate of 85%. Furthermore, recommendations for vaccinations should also vary within States. Our initial R0 estimates are driven mainly by “hotspots” within States, like New York City, where deaths increased most rapidly. More generally, we caution that vaccination programs must consider not only local infection rates but also movement of infected individuals between States (Roberts and Heesterbeek 2003). Nonetheless, the challenges of producing and delivering vaccines once they have been discovered calls for a hierarchical distribution program targeting those areas that have the highest potential R0.

These results highlight the importance of data availability and statistical tools for State-level decisions about public health interventions to manage the spread of COVID-19. Unfortunately, the results also highlight that States are apparently making decisions to relax public health interventions without considering evidence in the data.

## Data Availability

All code and data are provided in the Supporting Information. The data are from: The New York Times (2020) Coronavirus (Covid-19) data in the United States.

https://github.com/nytimes/covid-19-data

## Supporting Information

The Supporting Information consists of five sections: (A) Overall statistical rationale, (B) Estimating time-varying *r(t)*, (C) Data handling, (D) Assessment of robustness, and (E) Analysis of case data.

## Supporting Information

This technical supplement consists of five sections:

A. Overall statistical rationale
B. Estimating time-varying *r(t*)
C. Data handling
D. Assessment of robustness,
E. Analysis of case data

## (A) Overall statistical rationale

### 1. Estimating the rate of disease spread

The rate of spread of a disease in a population at the early phase of an epidemic, *r*_0_, when the entire population is susceptible depends on the basic reproduction number, R0, giving the number of secondary infections produced per infected individual, and the distribution of the time between primary and secondary infections. Thus, if the spread rate and distribution of infection times can be estimated, R0 can then be calculated. Our strategy is to estimate *r*_0_ as the most direct parameter associated with the dynamics of an epidemic, and then subsequently estimate R0.

The rate of spread of a disease can be estimated directly from the observed number of new deaths, or newly identified cases, in a population. The advantages of this approach to calculating *r*_0_, and from this R0, instead of case-tracking include: (i) it captures all of the real-life complexities that affect R0 by simply observing what happened in real life, and (ii) it uses data that are (tragically) becoming more prevalent. The challenges include (i) the changes in *r*(*t*) that are to be expected (and hoped for) as people and governments respond to lessen the spread, and (ii) the statistical challenges and uncertainties of determining rates of disease spread when the numbers of deaths and/or cases are still low.

We developed and tested statistical methods to overcome the two challenges of estimating R0 from death and case data. Because the rate of spread of a disease may change rapidly in response to actions that are taken to reduce disease transmission, we used a time-varying autoregressive model that allows for the rate of spread to change through time, *r(t*). Other models take a related approach. For example, the Imperial College of London model (Cori et al. 2013, Flaxman and al. 2020) assumes that the rate of spread changes immediately following actions taken by governments. Our approach differs, because we instead allow *r*(*t*) to change as inferred from the data: we do not make a priori assumptions about when these changes occur. Thus, we use an approach to analyzing time series in which the parameters of the time series model themselves can change through time, with the rate of change estimated from the data to give the best fit, as measured by the maximum likelihood. This is similar to the approaches taken by other researchers (Balabdaoui and Mohr 2020, Scire et al. 2020, Systrom and Vladeck 2020).

The second challenge is that the counts of deaths and cases at the beginning of an epidemic are low. To account for this, the time-series model includes increased uncertainty (measurement error) when the mean counts of deaths or cases are low. These low counts also mean that there is greater uncertainty in the estimates of *r(t*). Standard (asymptotic) approaches often have poor statistical properties (type I errors, correctly calculated confidence intervals) when sample sizes are small (Gelman and Hill 2007). Therefore, we use bootstrapping (Efron and Tibshirani 1993) in which simulation time series are reconstructed to share the same pattern as the observed time series; a large number of simulated time series are then fit using the same statistical model as used to fit the original data. This bootstrapping procedure thus gives estimates and confidence intervals for model fit to the real data.

Our approach focuses on estimating the time-varying rate of spread, *r*(*t*), of the number of deaths and reported cases of new infecteds. Some approaches explicitly model the rate at which an individual infects susceptibles as a probability function of the length of time the individual has been infected, and then model the probability of a newly infected individual being reported as infected (Cori et al. 2013, Thompson et al. 2019, Flaxman and al. 2020, Scire et al. 2020, Systrom and Vladeck 2020). In other words, these models are built around the processes of infection and reporting. We also built such a model, but we only use it to test the robustness of our statistical approach. For fitting the data, we instead use a simpler statistical model. Our rationale is that, for statistical fitting, it is better to keep the model as simple as possible, rather than “building in” assumptions about the processes of infection, reporting, and death. Our simple phenomenological model uses the same data as a more complicated, process-based model, and therefore both approaches ultimately rely on the same information. The simpler approach, however, does not depend on assumptions about the infection processes. Nonetheless, after fitting the simpler model additional assumptions can be made to interpret the fits. An advantage of the simple statistical model is that the assumptions about processes do not affect the results (fits), and how the assumptions affect the interpretation of the results is more transparent than if the assumptions are made in the statistically fit model.

The approach we applied to the data was the best of several approaches that we tried. We also performed analyses with a related model having time-varying *r(t*), but also allowing step-changes in *r(t*), with the location of the step changes selected to maximize the model likelihood. This approach, however, often led to model overfitting. Another approach we investigated was fitting a Generalized Additive Mixed Model (GAMM) to the death and case data while accounting for autocorrelation and greater measurement error at low counts (Wood 2017), and we tried a version of this GAMM model that allowed for step changes in *r*(*t*). However, the GAMM models tended to give greater bias than the approach we finally used. We fit the final models to the time series running both forwards and backwards to check that they gave consistent estimates.

After estimating *r(t)*, we computed R0(*t*) as 1/∑*_τ_*e^−^*^r(t^*^)^*^τ^p(τ*), where *τ* is the number days after initial infection, and *p(τ)* is the proportion of secondary infections produced per infected individual at *τ* (Dublin and Lotka 1925). This expression assumes that deaths (removal of individuals from the population) occur after all secondary infections have occurred. The statistical analyses focus on the estimates of *r(t*), and different distributions of *p*(*τ*) can be used as more information about the course of SARS-CoV-2 infection becomes available. We used the distribution of *p*(*τ*) from Li et al. (2020).

For death and case time series, we used data provided by the New York Times (2020).

### 2. Assessment of robustness

To assess the performance of our methods, we built an age-structured simulation SIR model (susceptible-infected-recovered), iterated on a daily time step to generate realistic data for which we know the true underlying infection rate and R0. We simulated time series under different scenarios of changes in infection rates and assessed whether the time-varying autoregressive model could correctly estimate the initial R0 at the start of the simulated epidemic. This gives an assessment of the robustness of the statistical model to realistic challenges in the data. The SIR model was parameterized using results from previous studies on data from the epidemic in Wuhan, China (Ferretti et al. 2020, Li et al. 2020).

### 3. Data selection and handling

For analyzing time series of death and case data, decisions have to be made about which data to use and when to start the time series for analyses. Case data depend on both the level of infection in a State and the (changing) proportion of infections that are reported. The latter depends on changes in public awareness, changes in criteria used for identifying cases of COVID-19, changes in availability of physicians and testing to diagnose cases, etc. These factors introduce not only uncertainty (measurement error) in case data, but also possible bias. For example, as the epidemic and public awareness spread, the proportion of infections that are reported might increase. This would lead to an artificially high estimate of the spread rate. Therefore, for estimating the spread rate early in the epidemic, we used daily death counts that are less prone to these types of biases. Indeed, using case data early in the epidemics gives somewhat higher estimates of R0 than using death data for almost all States (section (E) Analysis of case data).

Deciding when to initiate the time series for analysis involves balancing three factors. First, because our goal was to try to estimate R0 for a “naive” population in which few interventions were taken, pushing the initiation of the time series as early as possible is important. Second, initiating the time series earlier also means that the count data are sparser, increasing uncertainty in the estimates. Third, we wanted the R0 estimate to reflect conditions within a State and therefore exclude deaths or cases caused by infections contracted elsewhere and brought into the State. To balance these factors, we selected a threshold of 3 deaths per day, or 30 cases per day, as the starting point of the time series we analyzed. We determined when these thresholds were met using the GAMM in the R package ‘mgcv’ (Wood 2019) to smooth the time series. The choice of start threshold did not affect the estimate of R0 for most States, with the exception of Wisconsin (section (C) Data handling).

## (B) Estimating time-varying *r(t)*

### 1. Time-varying autoregressive model

The time-varying autoregressive model that we applied to the COVID-19 death and case data is a variant of the TVIRI (time-varying intrinsic rate of increase) model presented in Bozzuto and Ives (2019), which is an implementation of time-varying autoregressive models (e.g., Zeng et al. 1998, Ives and Dakos 2012, Bragina et al. 2018) that is designed explicitly to estimate the rate of increase of a variable using non-Gaussian error terms. We assume in our analyses that the proportion of susceptible people relevant to the local epidemic is close to one, and therefore there is no decrease in the infection rate caused by a pool of individuals who were infected, recovered, and were then immune to further infection. Thus, the variant of the TVIRI model we used here does not include a density-dependent term that would account for decreases in the proportion of susceptibles in the population.

The general specification of the model is

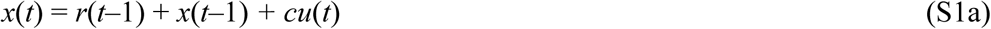

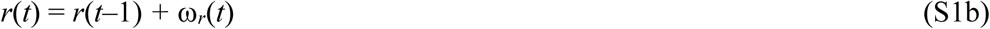

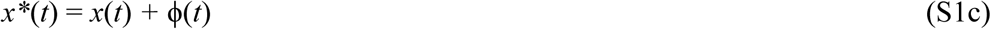

Here, *x(t*) is the unobserved, log-transformed value of deaths or cases at time *t*, and *x*(t*) is the observed count that depends on the observation uncertainty described by the random variable *ϕ*(*t*). Because a few of the datasets that we analyzed had zeros, we added 0.5 to the count data before log-transformation; other ways of treating zeros in the count data gave very similar results. The state variable *x*(*t*) can potentially depend on *u*(*t*), a measured environmental variable whose effect on *x*(*t*) is given by the coefficient *c*. This is used, for example, to produce a model in which there are break points in *r(t*). The model assumes that *x*(*t*) increases exponentially at rate *r*(*t*), where the latent state variable *r*(*t*) changes through time as a random walk with *ω_r_*(*t*) ~ N(0, σ^2^r). This assumption allows *r(t*) to change through time as dictated by the data, and the estimate of σ^2^r sets the rate at which *r(t*) can change from one day to the next.

We assume that the count data follow a quasi-Poisson distribution. Thus, the expectation of counts at time *t* is exp(*x(t*)), and the variance is proportional to this expectation. On the log-transformed scale of *x*(t)*, this implies that ϕ*(t*) has mean zero and variance approximately 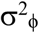 + exp(−*x(t*)), where 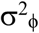 scales the variance.

We fit the model using the Kalman filter to compute the maximum likelihood (Harvey 1989, Durbin and Koopman 2012). In addition to the above-mentioned parameters σ^2^r, 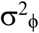, and *c* (if a covariate *u(t)* is included), we estimated the initial value of *r(t*) at the start of the time series, *r*_0_, and the initial value of *x(t*), *x*_0_. The estimation also requires an assumption for the variance in *x*_0_ and *r*_0_, which we assumed were zero and σ^2^r, respectively.

### 2. Interpretation of the time-varying autoregressive model

The time-varying autoregressive model estimates the rate of spread of the disease, *r*(*t*), from the count of deaths or new cases observed each day, *x*\*(*t*). Any value of *x*(t)* reflects the number of people infected over multiple days in the past, and the proportion that is counted as a result of dying or being diagnosed as infected on day *t*. If the disease had been spreading exponentially at constant rate for many days, and if the number of infected people was large, then the increase from *x*\*(*t−*1) to *x*(t)* would approach a constant value; in other words, *r(t*) would give the exponential rate of spread of the disease. This would be true even if only a small fraction of the infected population died or was diagnosed, provided these fractions did not change through time. However, changes in the infection rate will mean that the disease is not at its “stable infection age distribution”, the distribution of time since infection observed in the infected population (Caswell 1989). While this does not affect the statistical model fitting, it will mean that the observed spread of the disease is not exactly equal to the rate of new infections. Nonetheless, because the distribution of times between infection and counting (deaths or cases) is fairly broad, the assumption that populations are at their “stable infection age distribution” is unlikely to cause a great difference between the observed rate of disease spread and the infection rate. This is address in detail in the section (D) Assessment of robustness.

The “true” value of the number of daily deaths or new cases in the model, *x(t)* (S1a), is the probability that an infected or deceased person is counted. After accounting for measurement error (S1c), all of the variation in *x(t*) is assumed to be given by variation in the spread rate *r(t*) (S1b). Therefore, the variation ω*_r_(t*) in *r(t*) includes both the day-to-day variation in the spread rate and the longer-term changes in *r(t*) that results when estimates of ω*_r_(t*) have a mean different from zero. The assumption that *r(t*) is a random walk gives it flexibility to track the patterns in the data as the model is fit. We suspect that the true changes in the infection rate do not vary greatly on a day-to-day basis. This might argue for fitting a smoothing curve to *r*(*t*) or *x(t*); indeed, this is what the GAMM we used does. Nonetheless, we found that results from curve-fitting models were sensitive to decisions made about the type of curves that were fit. The time-varying autoregressive model was less dependent on a priori assumptions, due to few a priori assumptions about the data. Further, the bootstrapping method we applied to obtain estimates of the uncertainty of the model fits also acts as a smoothing method.

We derived estimates of R0(*t*) directly from *r(t*) as

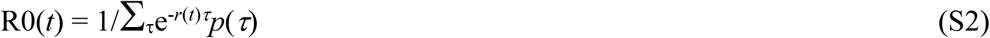

where *p*(*τ*) is the distribution of the proportion of secondary infections caused when by a primary infection that occurred *τ* days previously. We used the distribution of *p(τ)* from Li et al. (2020) that had an average serial interval of T0 = 7.5 days; smaller or larger values of T0, and greater or lesser variance in *p(τ)*, will decrease or increase R0(*t*) but will not change the pattern in R0(*t*) through time. We report values of R0(*t*) at dates that are offset by the average length of time between initial infection and death, or, for case data, the time between initial infection and reporting. These are taken as 18 days (Zhou et al. 2020) and 8 days (the serial interval Li et al. 2020), respectively. Thus, our statistical model uses only the raw data on the observed changes in deaths or cases to calculate *r*(*t*), rather than making assumptions about the timing of transmission, death, and reporting. Assumptions about timing are used only after the model fitting to calculate R0(*t*) and the timing of infections.

### 3. Parametric bootstrapping for uncertainty

To generate approximate confidence intervals for the time-varying estimates of *r*(*t*), we used a parametric bootstrap designed to simulate datasets with the same characteristics as the real data that are then refit using the autoregressive model. This procedure answers the question: If it were possible to observe many times series generated by the same process, how variable would be the results of the statistical model fit? This bootstrapping approach requires assumptions about the process underlying the true data. Because the underlying changes in *r(t*) are of interest, the bootstrap incorporates the time-varying changes in the estimated values of *r*(*t*) from the fitted data.

Changes in *r(t*) consist of unbiased day-to-day variation and the biased deviations that lead to longer-term changes in *r(t*). The bootstrap treats the day-to-day variation as a random variable while preserving the biased deviations that generate longer-term changes in *r(t*). Specifically, the bootstrap was performed by calculating the differences between successive estimates of *r(t*), ∆*r(t*) = *r(t) − r(t*-1), and then standardizing to remove the bias, ∆*r_s_(t)* = ∆*r(t*) − E[∆*r(t*)]. The sequence ∆*r_s_(t)* was fit using a autoregressive time-series model with time lag 1, AR(1), to preserve any shorter-term autocorrelation in the data, and then for the bootstrap a new time series was simulated from this AR(1) model, ∆*ρ(t*), and then standardized, ∆*ρ_s_(t)* = ∆*ρ(t)* − E[∆*ρ*(*t*)]. The simulated time series for the spread rate was constructed as *ρ(t)* = *r(t*) + *∆ρ_s_(t)/*2^1/2^, where dividing by 2^1/2^ accounts for the fact that ∆*ρ_s_(t)* was calculated from the difference between successive values of *r(t*). A new time series of count data, *ξ(t)*, was then generated using equation (S1a) with the parameters from fitting the data. Finally, the statistical model was fit to the reconstructed *ξ(t*). In this refitting, we fixed the variance in *r(t*), σ^2^r, to the same value as estimated from the data. Therefore, the bootstrap confidence intervals are conditional of the estimate of σ^2^r. We imposed this condition, because the estimate of σ^2^r tends to absorb to zero when the change in *r(t*) is small, with variation in *x(t*) transferred to the measurement error variance.

### 4. Example fits

As an example of the model fit, we will use death data for Wisconsin. We selected Wisconsin, because it posed the greatest challenge when deciding how to handle data (section (C) Data handling).

The number of deaths per day in Wisconsin increased rapidly before beginning to plateau (Fig. S1A). The differences between the fitted and observed death counts, *x(t*) and *x*(t*), are determined by measurement error. The time-varying estimates of *r(t*) decrease to near zero (Fig. S1B). The dashed lines in Fig. S1B give the standard deviations for the point estimates of *r(t*) produced by the Kalman filter. There is considerable day-to-day variation in the estimates of *r(t*), as anticipated by the observed changes in the death count. The bootstrap smooths out these fluctuations (Fig. S1C). Furthermore, when the fitted line strays from the bootstrap confidence intervals, the data changes in a manner that cannot be captured by the bootstrap procedure. In other words, the deviation in *r*(*t*) estimated from the data exceeds that expected from model underlying the bootstrap. Because the bootstrap assumes that the variance in *r(t*) is homogeneous over the time series, places where *r(t*) strays from the bootstrap suggest more-rapid changes in the estimates of *r(t*) than elsewhere in the time series. Thus, the bootstrap gives a way to assess lack-of-fit of the data to the assumptions of the model.

### 5. Fitting reversed time series

A useful model diagnostic is to fit the model to the reverse of a time series to determine whether the fit changes. We did this with two goals. First, time-series models require assumptions about the variances at the start of the time series, in our case the variances in *r*_0_ and *x*_0_. Reversing the time series investigates this, because the variances of *r*_0_ and *x*_0_ in the reversed time series will have variances that have been accumulated over the future values that were reversed. Second, the model (S1a-c) updates changes in *r(t*) from changes in *x(t*), and if there are rapid changes in the true value of *r(t*), changes in the estimates of *r(t*) will occur with a lag. This lag can be captured by reversing the time series.

Fitting the reversed time series shows little effect of initial assumptions about the variances in *r*_0_ and *x*_0_, because the estimates of *r*_0_ are the same in the forward and reversed analyses (Fig. S2). Nonetheless, the analysis of the reversed time series showed an earlier decrease in *r(t*). This suggests a delayed response of the model fit to true changes in *r(t*). Despite this delay, however, estimates of *r*_0_ for forward and reverse time-series fitting were similar for all States (Fig. S3). The exception is Wisconsin, which is discussed in more detail in the section (C) Data handling.

### 6. Alternative statistical models

We investigated a variant of our approach in which we allowed *r(t*) to change as a step function, with up to two steps. The model was fit with all possible step-change time points that were separated by at least 6 days, with the best model selected as the one having the highest log likelihood. Thus, we did not discount for the number of parameters, although using AIC or other selection criteria is also possible. We then used the same bootstrap procedure to obtain confidence intervals. As the example for Wisconsin shows (Fig. S4), this often gave similar results to the time-varying model, although in some cases it gave wide confidence bounds that suggests overfitting.

As an alternative approach, we fit a GAMM to the daily death or case count data assuming ARMA(1,1) residuals and quasi-Poisson measurement error (Wood 2017, 2019). We then calculated the values of *r(t*) as the differences *x(t*) − *x(t*-1). The GAMM requires selecting the number of additive functions, and using AIC to select the model of appropriate complexity generally led to simple models that were too “stiff” to capture rapid changes in *r(t*). Increasing the number of functions, for example *k* = 10, led to more dynamic changes in *r(t*) (Fig. S5). Nonetheless, the bootstrapped confidence bounds indicate that the GAMM is prone to missing true changes in *r*(*t*).

## (C) Data Handling

### 1. Selecting the initial time point

The estimates of *r*_0_ depend on when the time series is started, that is, what date (Julian day) is regarded as time zero. We selected a threshold of 3 deaths per day, or 30 cases per day, as the starting point of the time series we analyzed. We determined when these thresholds were met using the GAMM (Wood 2019) to smooth the time series. This avoids the bias that would be caused if a criterion such as “the first day on which deaths exceed 3” were used.

Wisconsin proved to be a complicated case for which small changes in the threshold used to start the time series made a large difference in the estimate of *r*_0_. This appears to be due to a single zero early in the epidemic. When we used a threshold of two rather than three days, the estimate of *r*_0_ dropped from 0.28 to 0.12 (Fig. S6). This also brings the bootstrap estimates of *r*_0_ from analyses of the forward and reversed time series into agreement (Fig. S3).

### 2. Weekly artifacts in reporting

In death and case data aggregated for the entire USA, weekly cycles begin to appear in late April and May (New York Times 2020) with low numbers of cases occurring on Sunday and Monday. We suspect that this is due to reporting bias. An alternative explanation is that it is a rippling effect caused by the serial interval of roughly a week, so that a burst of infection in one week will generate a secondary burst the following week. However, because the data are aggregated for the entire USA, it is unlikely that these bursts would be synchronized sufficiently to generate the observed weekly cycles in the data. For the State data, however, these cycles are rarely observed and do not affect the analyses, except for the case data in Michigan, New York, Ohio, and South Dakota. These cycles were never observed in the early parts of the time series, and therefore our estimates of *r*_0_ should not be affected from the death data.

## (D) Assessment of Robustness

To assess the robustness of the statistical model, we built a simulation SIR model (susceptible-infected-recovered) of a hypothetical epidemic. The simulation model was not the same as the statistical model, so the goal was to determine whether the phenomenological statistical model was capable of capturing the rate of infection spread in the process-based simulations.

The simulation model tracks the number of infected individuals on day *t* who were infected *τ* days previously, *X(t;τ*). After 25 days, they are all assumed to be recovered or died. The probability distribution of the day on which a susceptible is infected, *p(t*), is given by a Weibull distribution with mean 7.5 days and standard deviation 3.4 (Li et al. 2020) (Fig. S7A). For an individual who dies, the day of death, *d(t*), is given by a Weibull distribution with mean 18.5 days and standard deviation 3.4 (Li et al. 2020) (Fig. S7B). Finally, for case data we need to know the time between initial infection and diagnosis, *h(t*), which we assume is lognormally distributed with mean 5.5 days and standard deviation 2.2 (Ferretti et al. 2020) (Fig. S7C).

On day *t*, the number of new infections produced by individuals who were infected *t* days earlier is *b(t*)*p(τ)*. The term *b(t*) is closely related to R0(*t*), the number of secondary infections caused per infection. However, because we allow *b*(*t*) to fluctuate on a daily basis, here we use a notation that differs from R0(*t*). Note, however, that on average R0(*t*) = ∑*_τ_ b*(*t* + *τ*)*p*(*τ*). The total number of new infections on day *t* is given by a lognormal Poisson distribution in which the mean of the Poisson process is *b*(*t*) *α*(*t*) ∑*_τ_ p*(*τ*)*X(t*; *τ*), where the lognormal random variable *α(t*) is included to represent environmental variation.

Deaths occur according to a binomial distribution for each infection age category *X(t*; *τ*), so that the probability of death of individuals that had been infected *τ* days earlier is (1 − *s*) β(*t*) *d*(*τ*), where *s* is the overall survival probability and β(*t*) is a lognormal distribution. We assume that the overall survival probability for COVID-19 is 98%; changes in this assumption had little effect on the simulation study. Once an individual dies, they are removed from the pool of individuals.

Cases are reported according to a binomial distribution for each infection age category *X(t*; *τ*), so that the probability of a person with the infection for *τ* days being diagnosed is *G* χ(*t*) *h*(*τ*), where *G* is the overall probability that a case is reported, and χ(*t*) is a logit-normal distribution to represent daily variation in reporting. We assume that the overall reporting probability is *G* = 0.5.

To illustrate the simulations, we assumed that the expectation of the infection rate, *b(t*), changes as a step function (Fig. S8A, black line), while there is also daily variation around this expectation (Fig. S8A, points). We also calculated the asymptotic rate of disease spread, R0(*t*) (Fig. S8A, red line). This shows that the expected daily infection rate, *b*(*t*), is closely related to the population-level R0(*t*). Over the simulated time series of 60 days, we then recorded the number of deaths (Fig. S8B) and diagnosed cases (Fig. S8C). We initiated the simulation with a single cohort of individuals, all infected on day 1 (Fig. S8C, filled black dot). This gives the “worst-case” situation in which the distribution of times-since-infection is far from the stable age distribution.

We fit this simulated dataset using the same procedure as we used for the real data, including the same rules to determine which day to initiate the fitted time series (Fig. S9). To compare the model fit of *r*(*t*) to the simulation, we computed the asymptotic rate of spread for the simulation model (Fig. S9B, red line). In this example, the statistical model gave a reasonable fit to the simulated data. We do not expect a perfect fit, because the simulation model had a step change in *r(t*) that cannot occur in the statistical model.

We performed a similar exercise while assuming that the expectation of the infection rate, *b(t*) changes geometrically, producing a linear change in *r(t*) (Fig. S10). In this particular example, the estimated values of *r*(*t*) are below the true values in the simulation in the first part of the time series. This downward bias in the early estimates of *r(t*) was not uncommon.

We performed 100 simulations with the expectation of *b(t*) changing as either a step function (Fig. S11) or geometrically (Fig. S12) to assess the overall robustness of the modeling approach. We investigated bias in the estimates of *r*_0_ (e.g., black line in Figs S9B and S10B) by generating a histogram of the differences between the estimated and true value of *r*_0_ from the simulation model, where the estimates were made using both the forward and reversed time series, respectively (Fig. S11). The estimates of *r*_0_ for the simulation model with step changes in *r(t*) (e.g. Fig. S9) were unbiased, whereas the estimates for the simulation model with geometric changes (e.g., Fig S10) were biased downwards (underestimates). We performed the same comparison using the mean of the bootstrapped estimates of *r*_0_ and obtained the same pattern and magnitude of bias (Fig. S12). For reporting the estimates of *r*_0_ and R0, we used the mean of the bootstrapped estimates of *r*_0_, because they are necessarily centered between the bootstrapped confidence intervals. Finally, we also investigated bias in the estimates of *r*_0_ from simulated case data using mean of the bootstrapped values (Fig. S13) which showed similar patterns as the death data (Fig. S12), although the analyses of the reversed time series with step changes in *r*(*t*) showed some positive bias.

We further used the simulations to assess the accuracy of the bootstrapped confidence intervals. Because the estimates of *r*_0_ for the simulation with geometric changes in *r*(*t*) were biased, and we wanted to know whether the width of the confidence intervals were accurate, we counted the number of simulations in which the true value of *r*_0_ lies outside the confidence bounds after subtracting the observed bias (Fig. S12). The tendency for the method to underestimate *r*_0_ is apparent in the bias-corrected 95% confidence intervals (Table S1) that are too low on the upper end, leading to more true values of *r*_0_ exceeding the confidence bound than expected.

In summary, the simulation study shows that the approach is reasonably robust when applied to realistic data. When the true *r*(*t*) changes slowly and continuously, the approach gives estimates that are downward biased, although there was little bias in the estimates when *r*(*t*) showed steps changes (Figs. S12, S13). Furthermore, even with the bias, the width of the confidence intervals for *r*_0_ was fairly accurate, although the upper confidence bounds were a little narrow.

### (E) Analysis of case data

We analyzed the time series of reported cases of COVID-19 using the same approach as we did with the death data. For most States, the estimate of *r*_0_ from the case data was slightly higher than for the death data (Fig. S14). The fits for Michigan and Virginia were visually poor (e.g., Fig. S15); in both cases, there were patterns in the latter part of the time series that forced a high estimate of σ^2^r and a very imprecise estimate of *r*_0_. A simple solution is to discard the latter part of the time series. However, we do not pursue this here and instead use this example to show that the method can fail when there are clear confounding effects in the data.

Also, for the estimate of *r*_0_ in Wisconsin (Fig. S14) we used an initial threshold of two deaths per day, rather than three, as described above (Fig. S6). This estimate, *r*_0_ = 0.12, is close to the estimate obtained from the case data (*r*_0_ = 0.10).

The higher estimates of *r*_0_ from case data relative to death data have at least two explanations. First, this pattern is consistent with greater reporting of infections as public awareness increased, leading to an observed increase in *r*_0_ due to changes in reporting bias. Second, the simulation study showed that the estimates of *r*_0_ from analyses of death data tended to be more downward biased than analyses of case data, presumably due to the lower numbers of deaths than cases. Because we could not distinguish these two explanations, we took the more conservative approach of using death data that led to lower estimates. Note that this is conservative from a statistical perspective, although for designing public health measures, it would be more conservative to use higher estimates of *r*_0_ and R0.

---

**Table S1.** Proportion of simulations in which lower and upper 66% and 95% confidence intervals were exceeded. The expected values are 0.025 for 95% confidence intervals and 0.17 for 66% confidence intervals.

| data | shape | lower 95% | lower 66% | upper 66% | upper 95% |
| --- | --- | --- | --- | --- | --- |
| deaths | step | 0.01 | 0.12 | 0.24 | 0.061 |
| deaths | geometric | 0.01 | 0.22 | 0.28 | 0.130 |
| cases | step | 0.04 | 0.18 | 0.20 | 0.050 |
| cases | geometric | 0.03 | 0.24 | 0.24 | 0.091 |

**Fig. S1.**
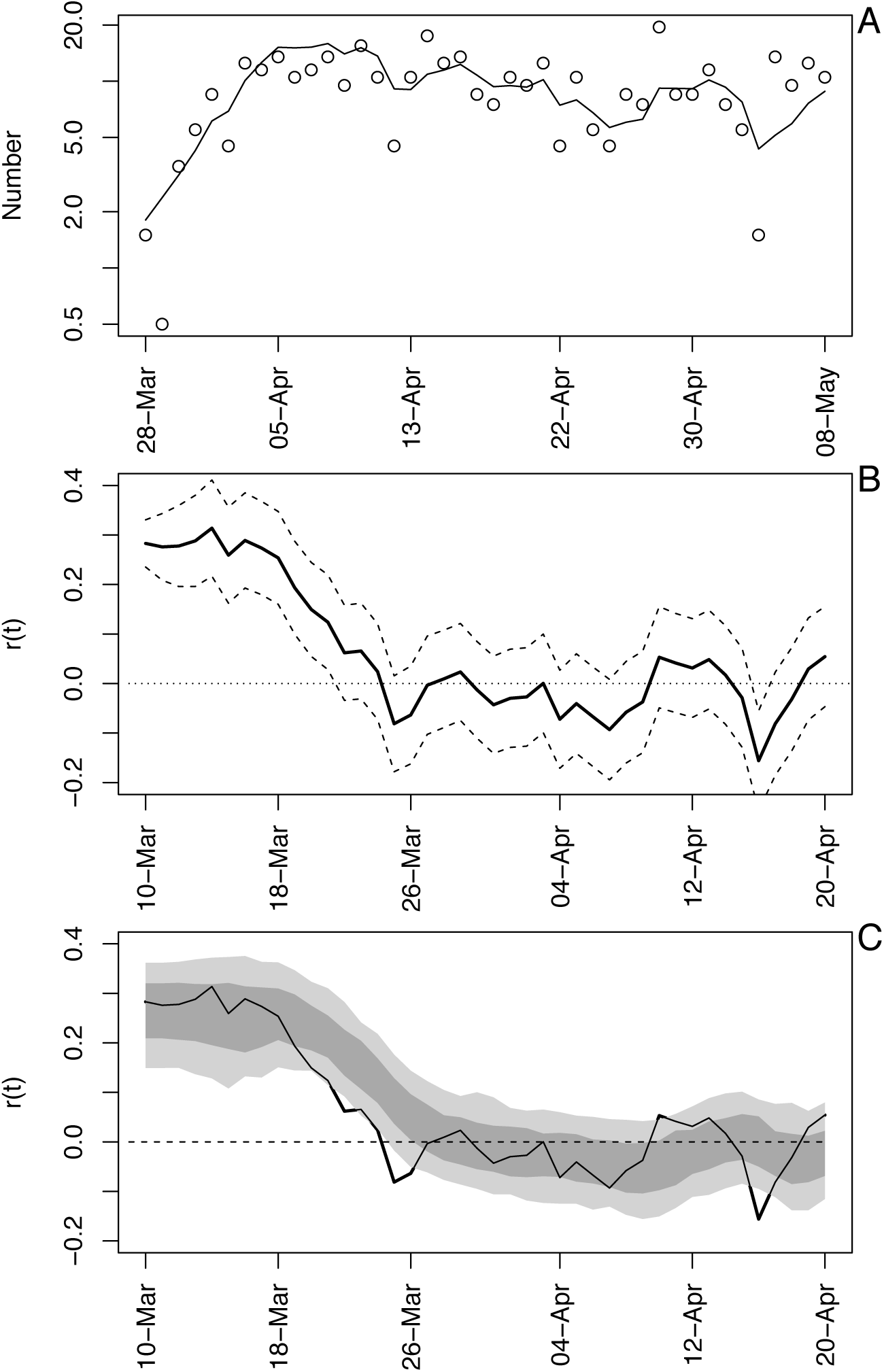
Time-varying state-space model fit to death data from Wisconsin. **(A)** Observed and fitted (line) deaths counted per day. **(B)** Estimated values of *r*(*t*) with ±1 SD from the statespace model (Eq. S1a-c). The dates have been offset by 18 days, the average time between infection and death. **(C)** Estimated values of *r*(*t*) with bootstrap 66% and 95% confidence intervals (dark and light gray, respectively) from 300 bootstrap simulations.

**Fig. S2.**
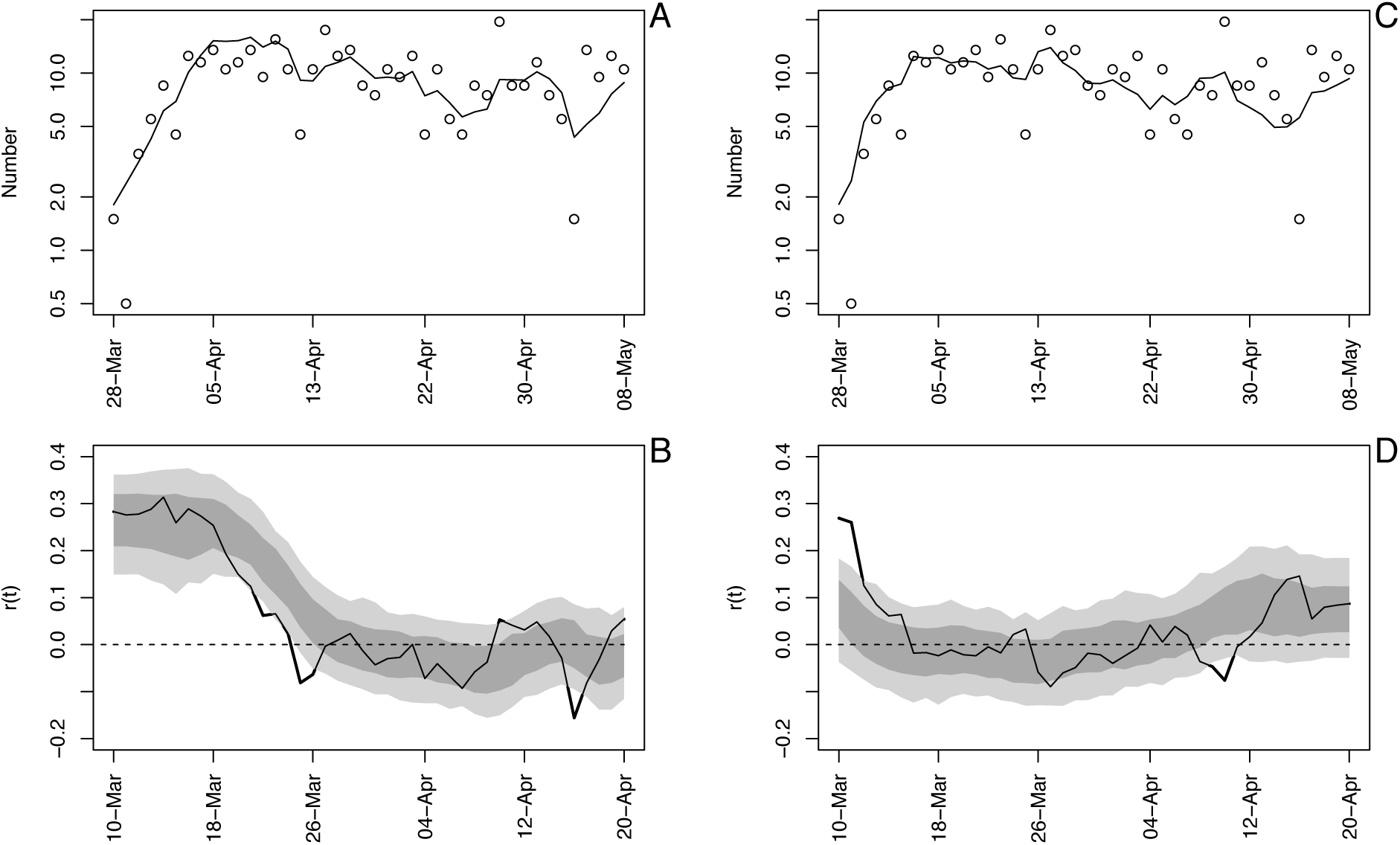
Time-varying state-space model fit to death data from Wisconsin using analyses in the forward **(A, B)** and backward **(C, D)** directions. See also figure S1.

**Fig. S3.**
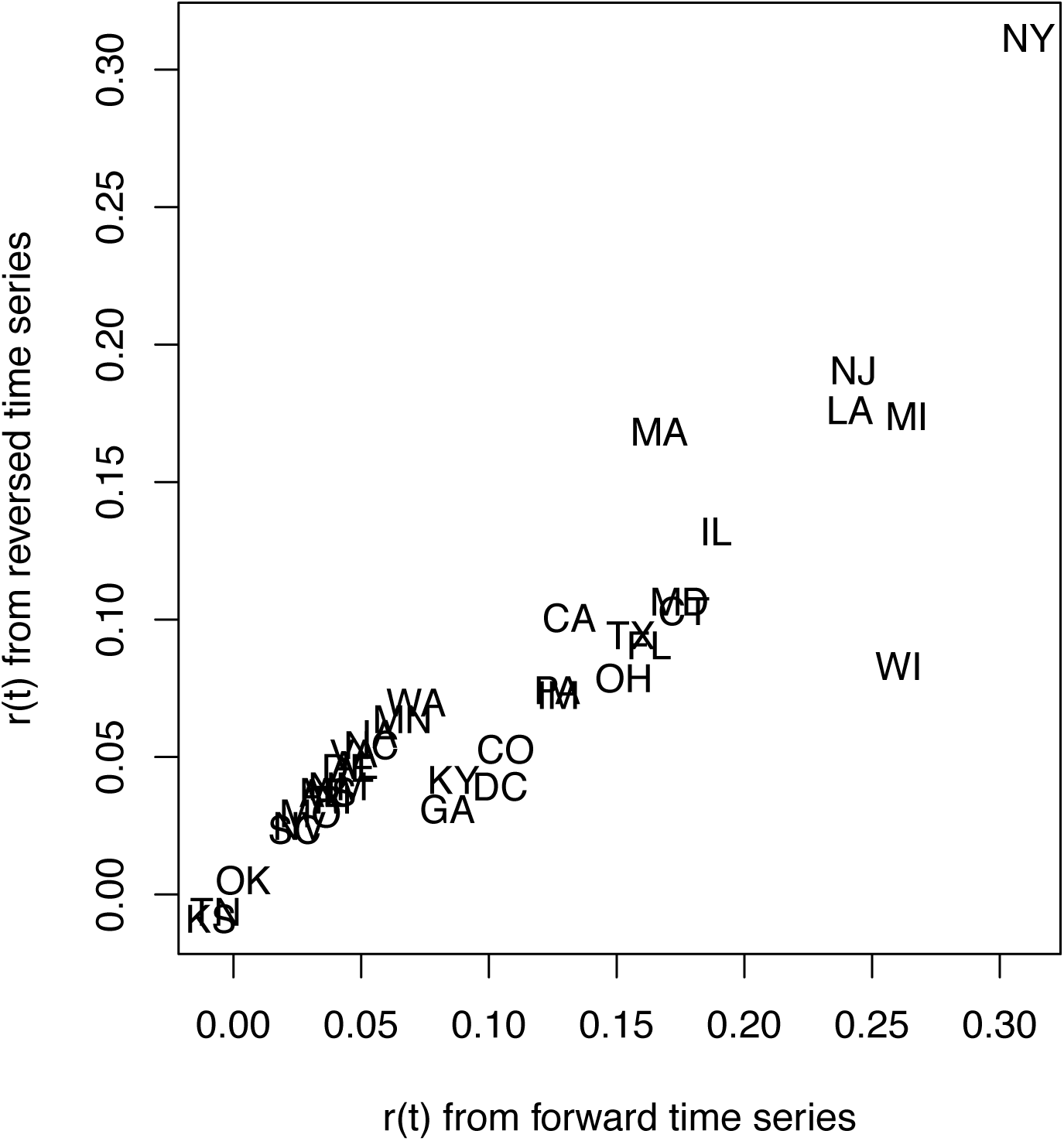
Estimates of the initial R0 for 36 States obtained by analyzing time series in the forward and backward directions.

**Fig. S4.**
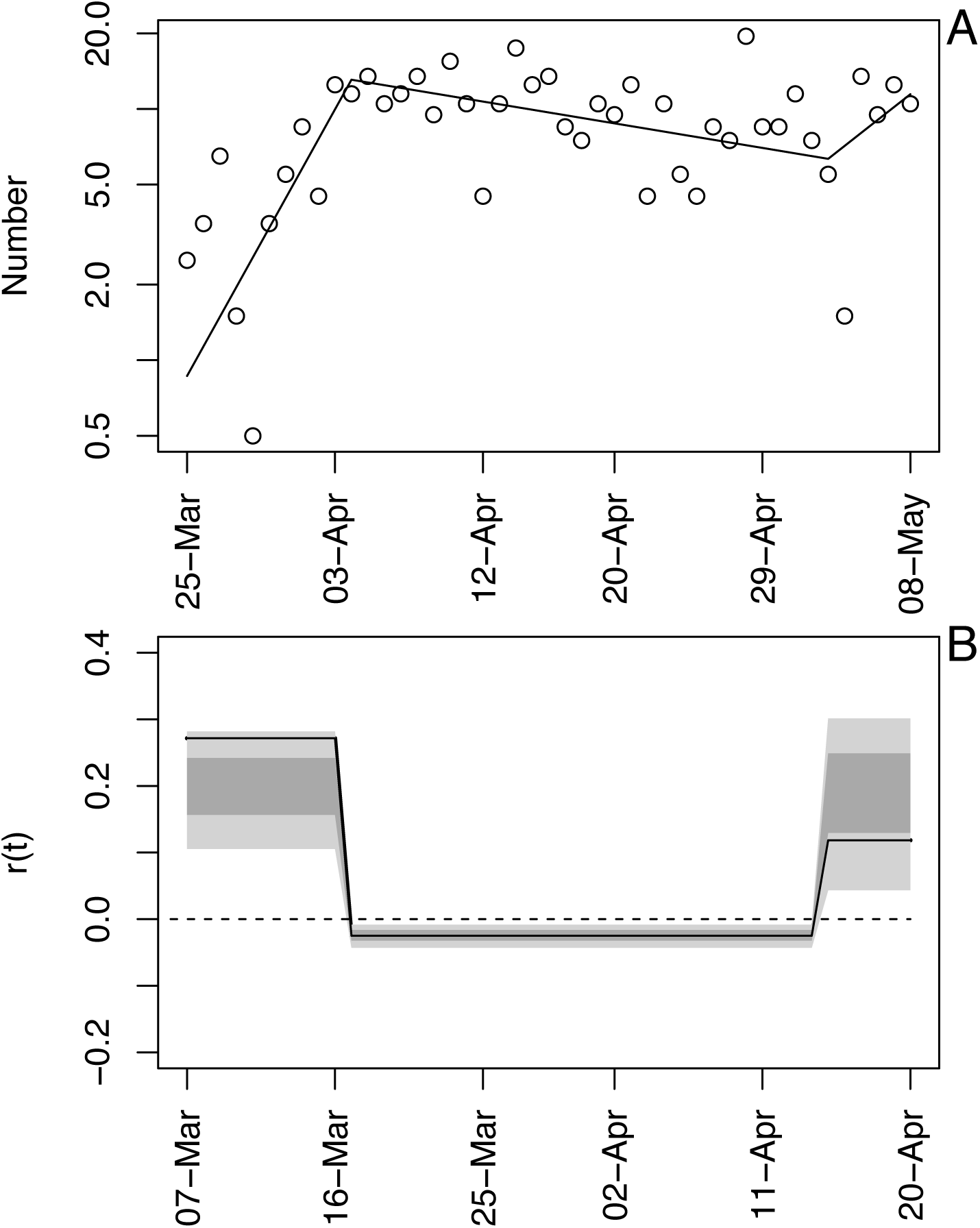
Time-varying state-space model fit to death data from Wisconsin in which up to two breakpoints are allowed. The locations of the breakpoints were selected to give the highest likelihood, conditional on breakpoints being at least 7 days apart.

**Fig. S5.**
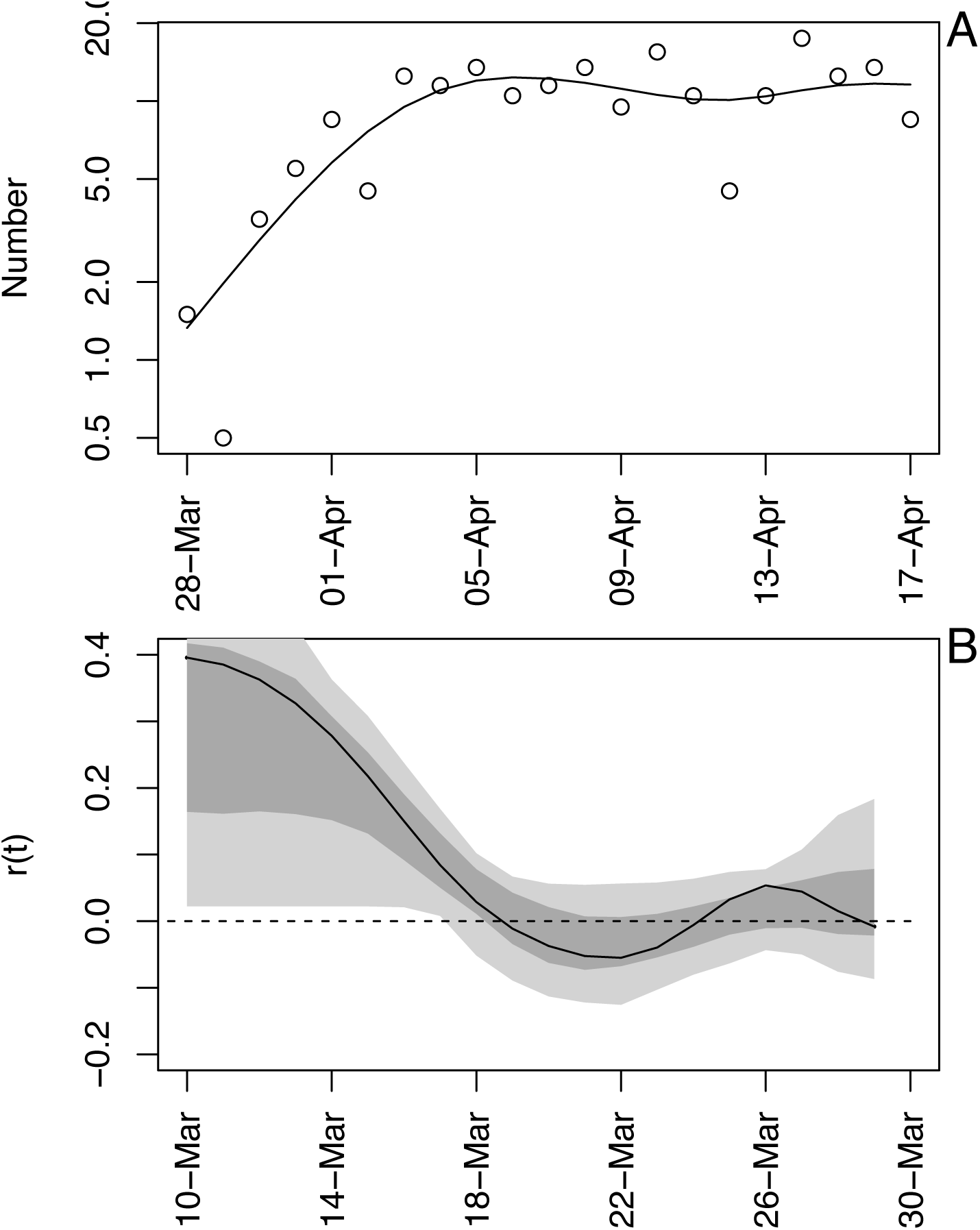
GAMM model fit to death data from Wisconsin. For the full data (Fig. 1), the GAMM fit a straight line to *x\**(*t*), and therefore for this example the time series was shortened by 21 days. **(A)** Observed and fitted (line) deaths counted per day. **(B)** Estimated values of *r*(*t*) with bootstrap 66% and 95% confidence intervals (dark and light gray, respectively). The bootstrap was performed by simulating data from the GAMM and re-fitting 300 simulated datasets. The GAMM was fit assuming *k* = 10 additive functions, ARMA(1,1) autocorrelated residuals, and variances of *x*(*t*) equal to exp(–*w x*(*t*)) where the coefficient *w* is estimated during the fitting process.

**Fig. S6.**
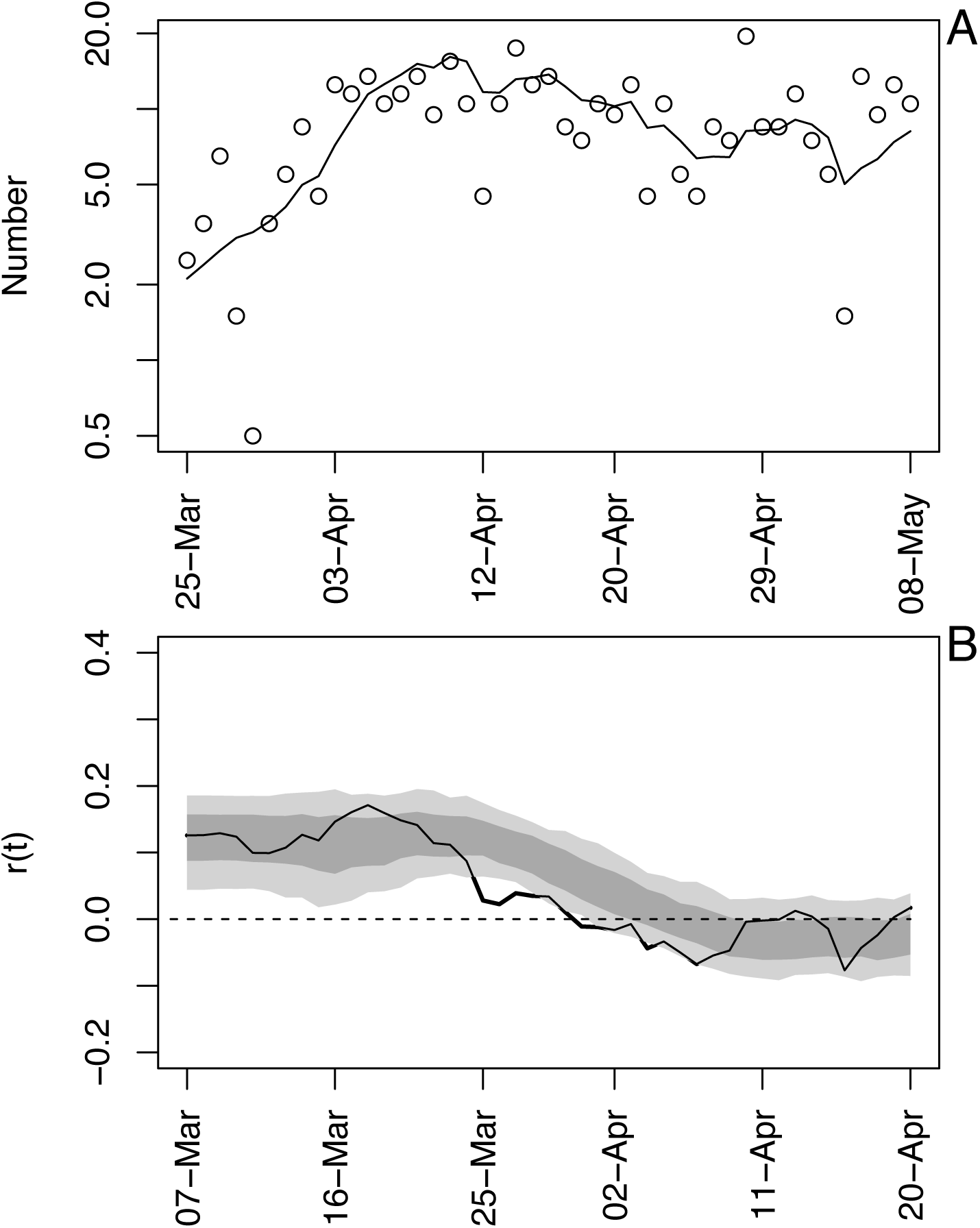
Time-varying state-space model fit to death data from Wisconsin. In contrast to Fig. S1, here the threshold to determine when to initiate the time series was set to two deaths per day, rather than three; all else is the same. **(A)** Observed and fitted (line) deaths counted per day. **(B)** Estimated values of *r*(*t*) with bootstrap 66% and 95% confidence intervals (dark and light gray, respectively).

**Fig. S7.**
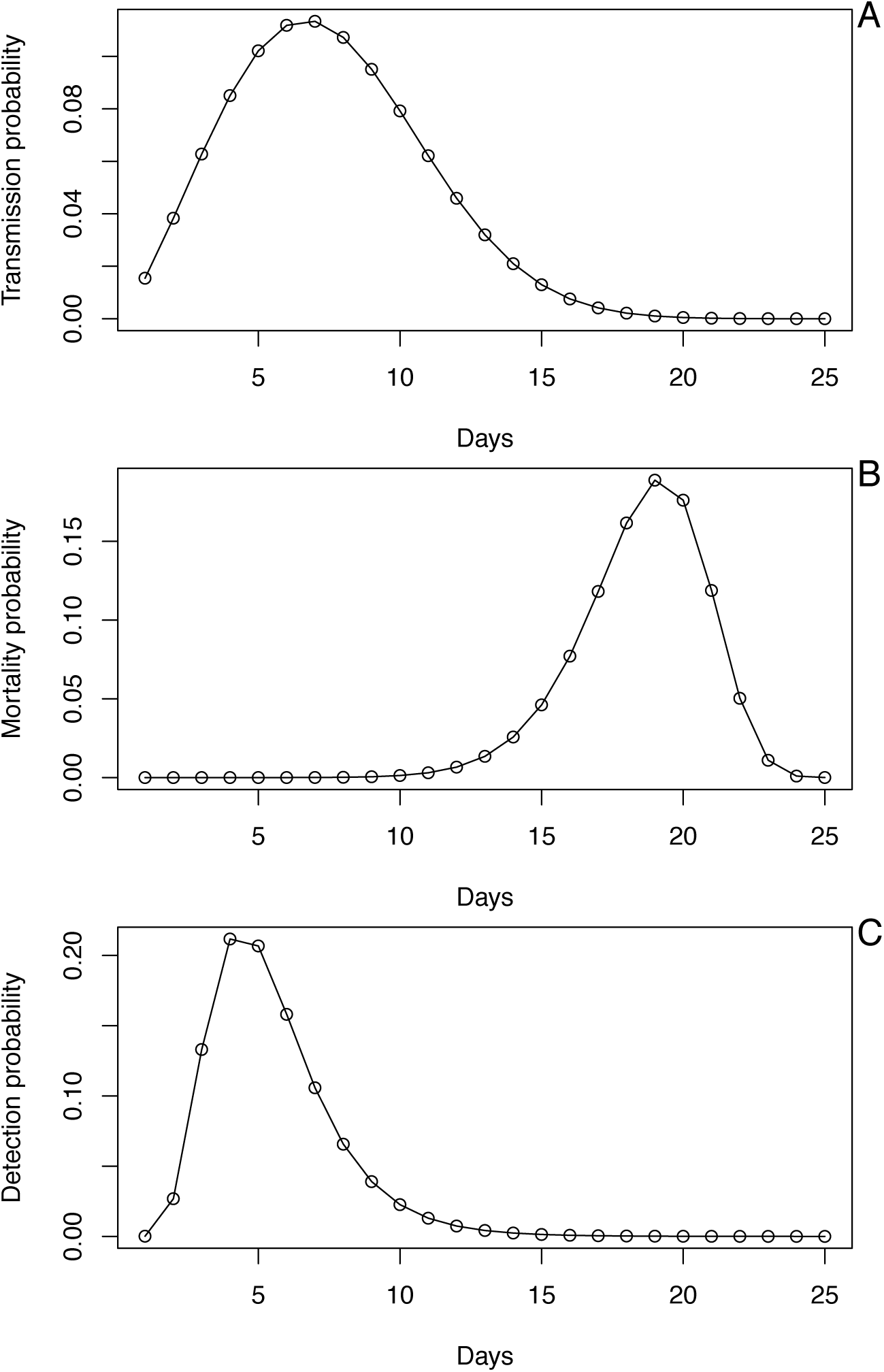
Probability distributions used in the process-based SIR simulation model used to test methods for robustness. **(A)** The probability distribution of the day on which a susceptible is infected, *p*(*t*), which is given by a Weibull distribution with mean 7.5 days and standard deviation 3.4. **(B)** For an individual who dies, the day of death, *d*(*t*), which is given by a Weibull distribution with mean 18.5 days and standard deviation 3.4. **(C)** For case data, the time between initial infection and diagnosis, *h*(*t*), which is lognormally distributed with mean 5.5 days and standard deviation 2.2.

**Fig. S8.**
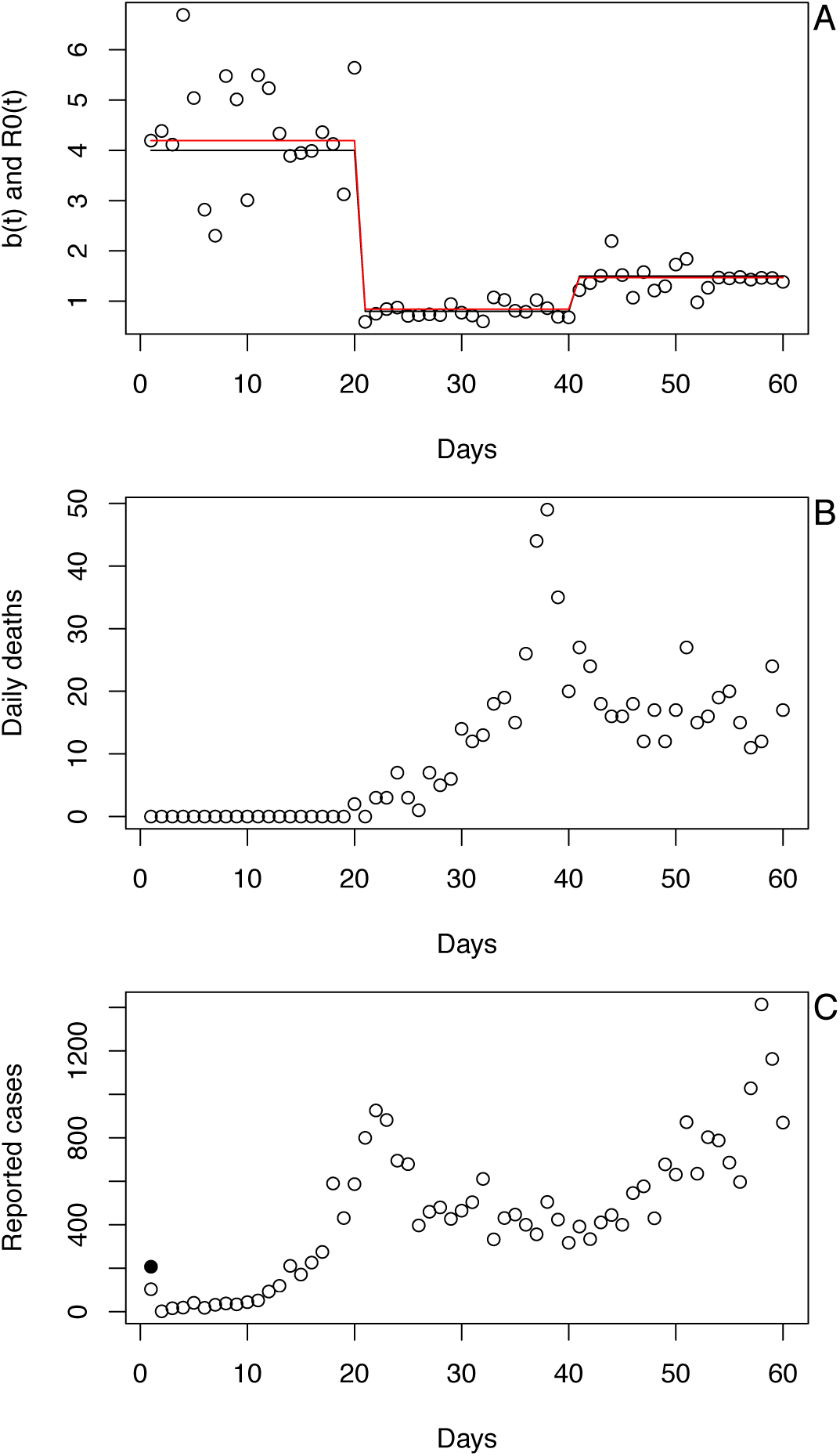
Example simulation from the process-based SIR model. **(A)** Changes in the infection rate, *b*(*t*), are modeled as a step function (black line) with daily variation (points). R0(*t*) (red line) tracks changes in *b*(*t*). **(B)** and **(C)** The number of deaths (B) and diagnosed cases (C) when the simulation is initiated with a single cohort of individuals, all infected on day 1 (solid black dot).

**Fig. S9.**
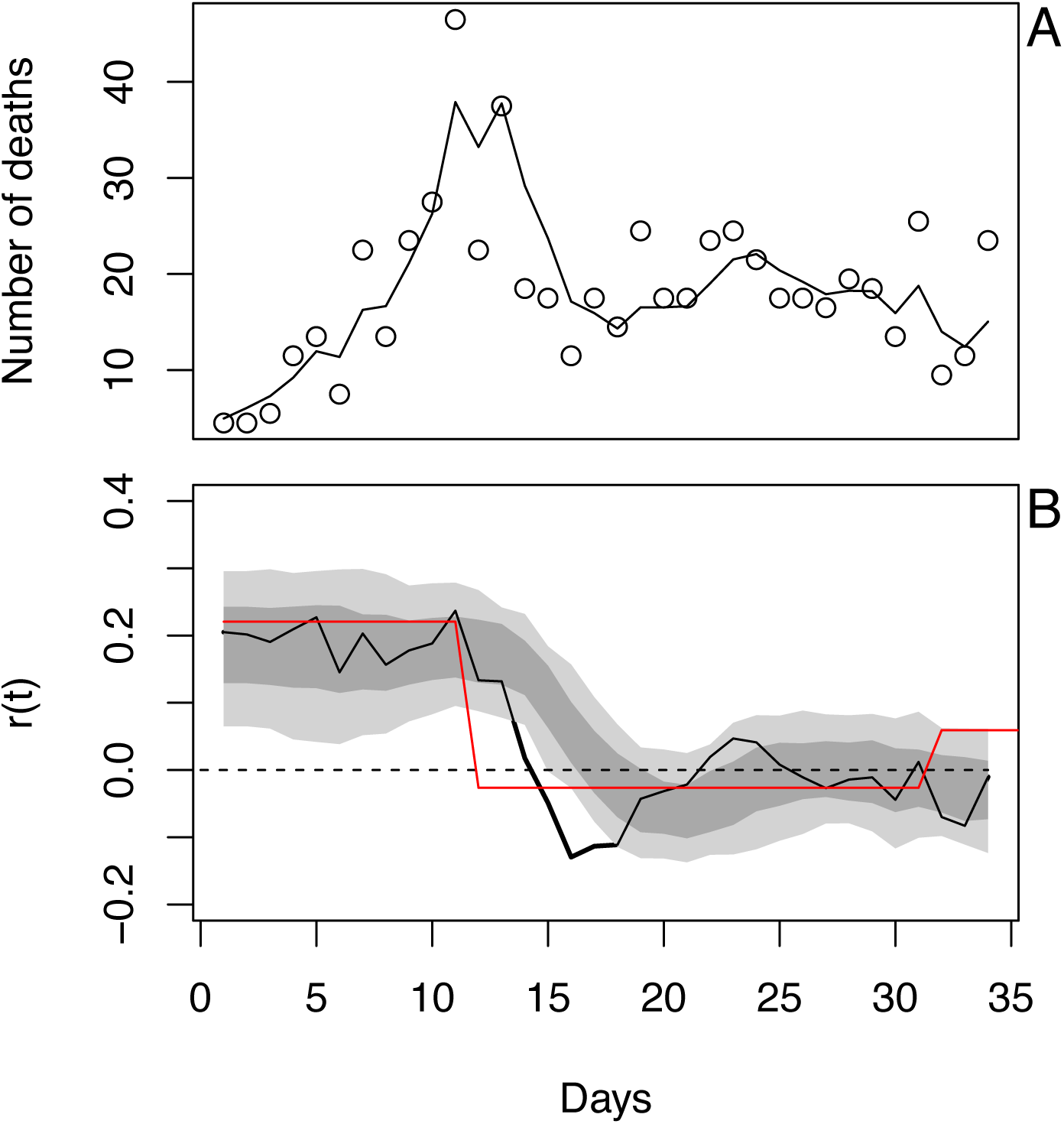
Statistical model (Eq. S1a-c) fit to simulated data with step changes in *b*(*t*) using the same procedure used for the real data (Fig. 1). In (B), the red line gives the asymptotic rate of spread calculated from the simulation model (i.e., ln(*λ*(*t*)), where *λ*(*t*) is the leading eigenvalue of the iterated age-of-infection structured matrix underlying the SIR).

**Fig. S10.**
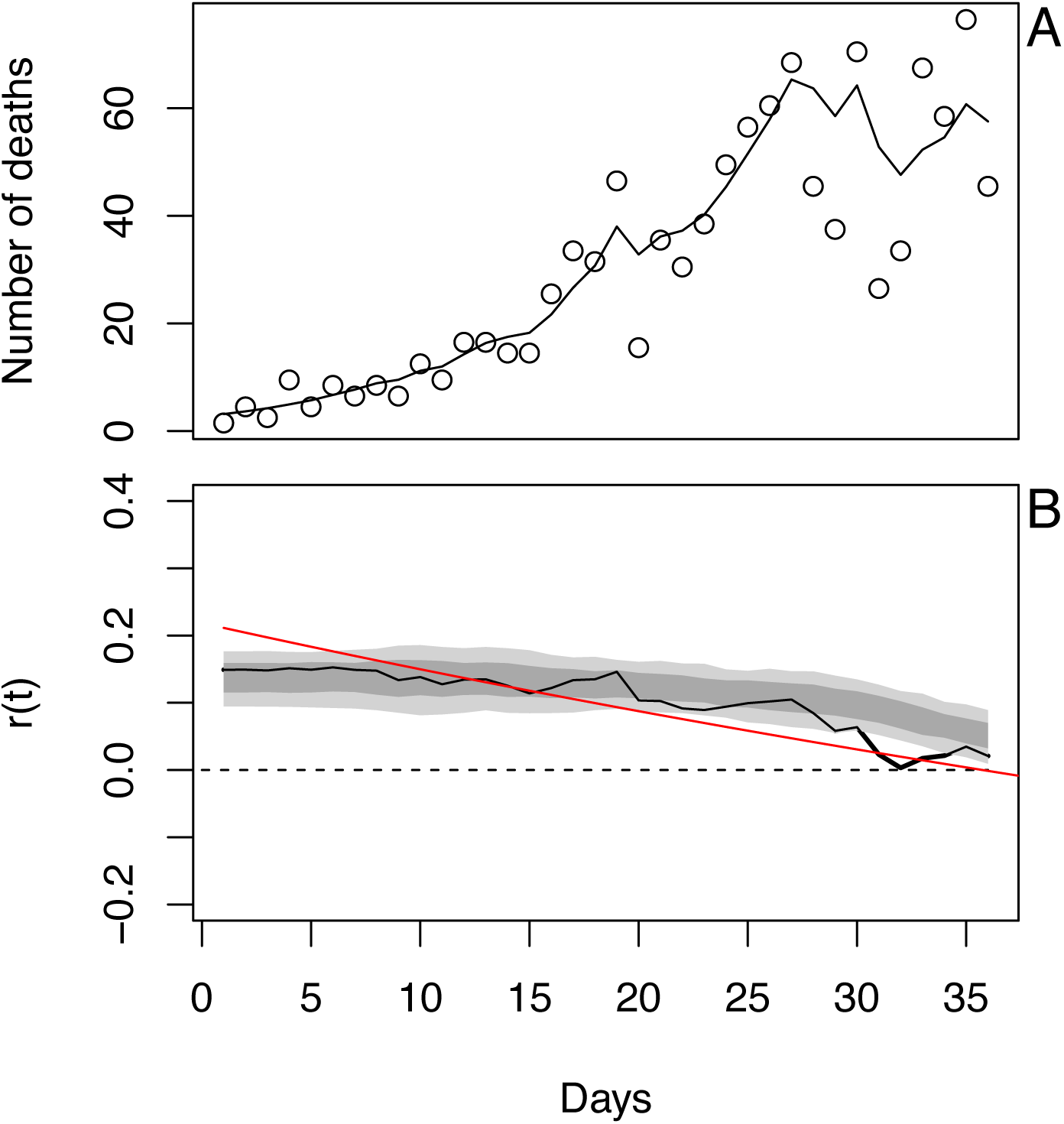
Statistical model (Eq. S1a-c) fit to simulated data with geometric changes in *b*(*t*). See figure S9.

**Fig. S11.**
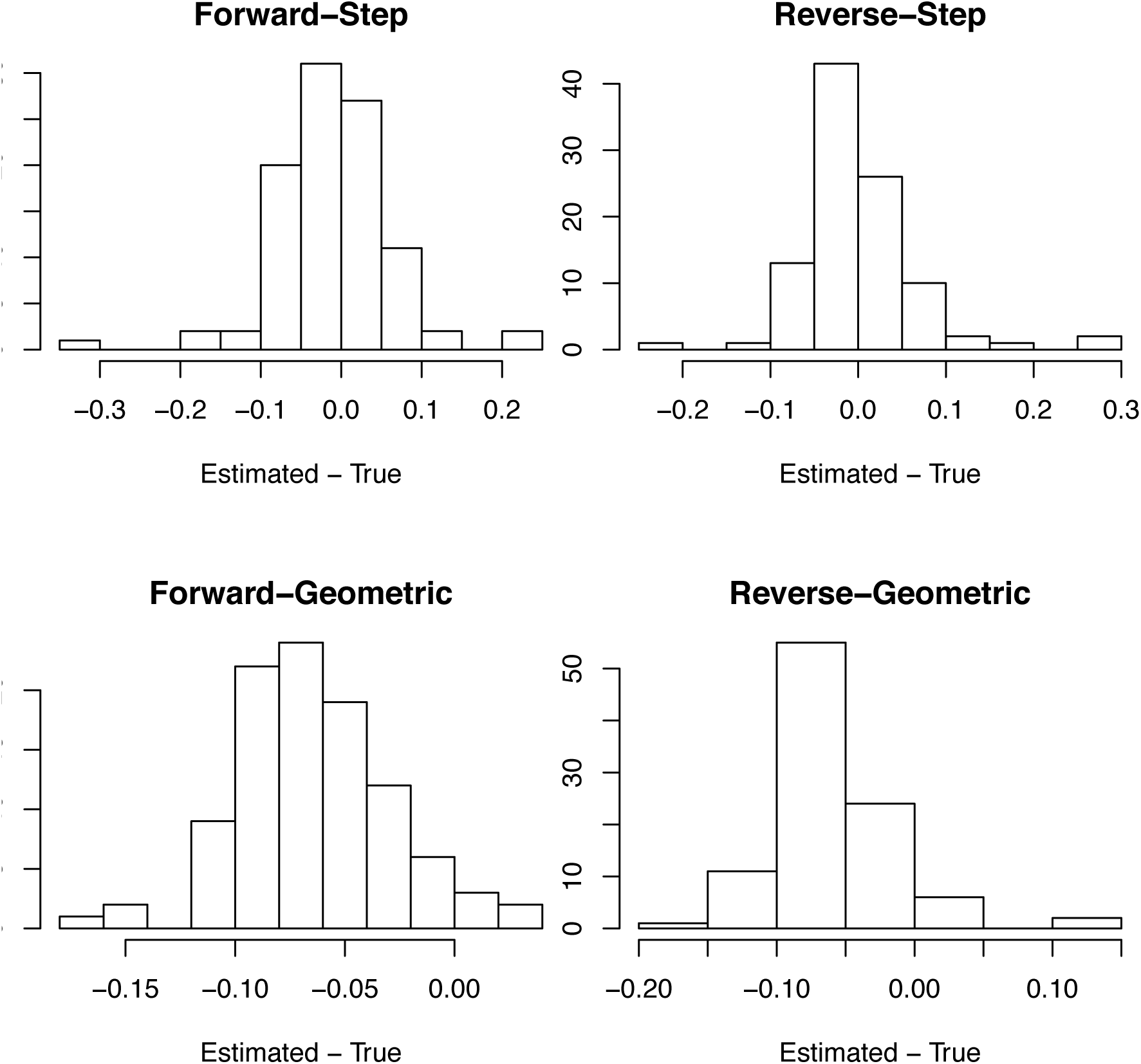
For death data, bias in the point estimates of *r*_0_ as the difference between the estimated and true values of *r*_0_ in the simulations. Simulations where performed in which *r*(*t*) experienced either step changes (Fig. S9) or geometric changes (Fig. S10), and the time series were either fit in the forward or reverse directions.

**Fig. S12.**
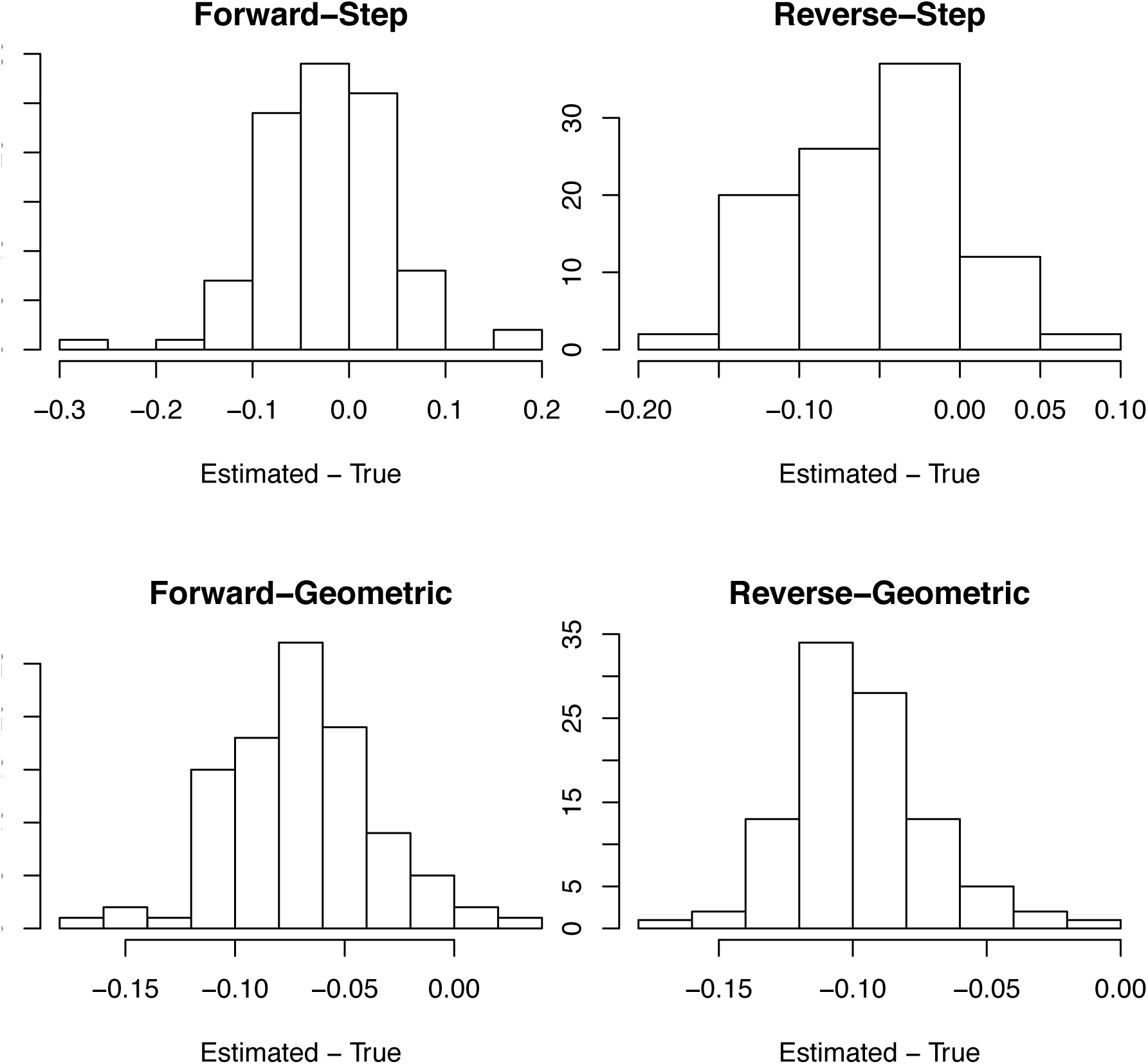
For death data, bias in the bootstrap estimates of *r*_0_ as the difference between the estimated and true values of *r*_0_ in the simulations. See figure S11.

**Fig. S13.**
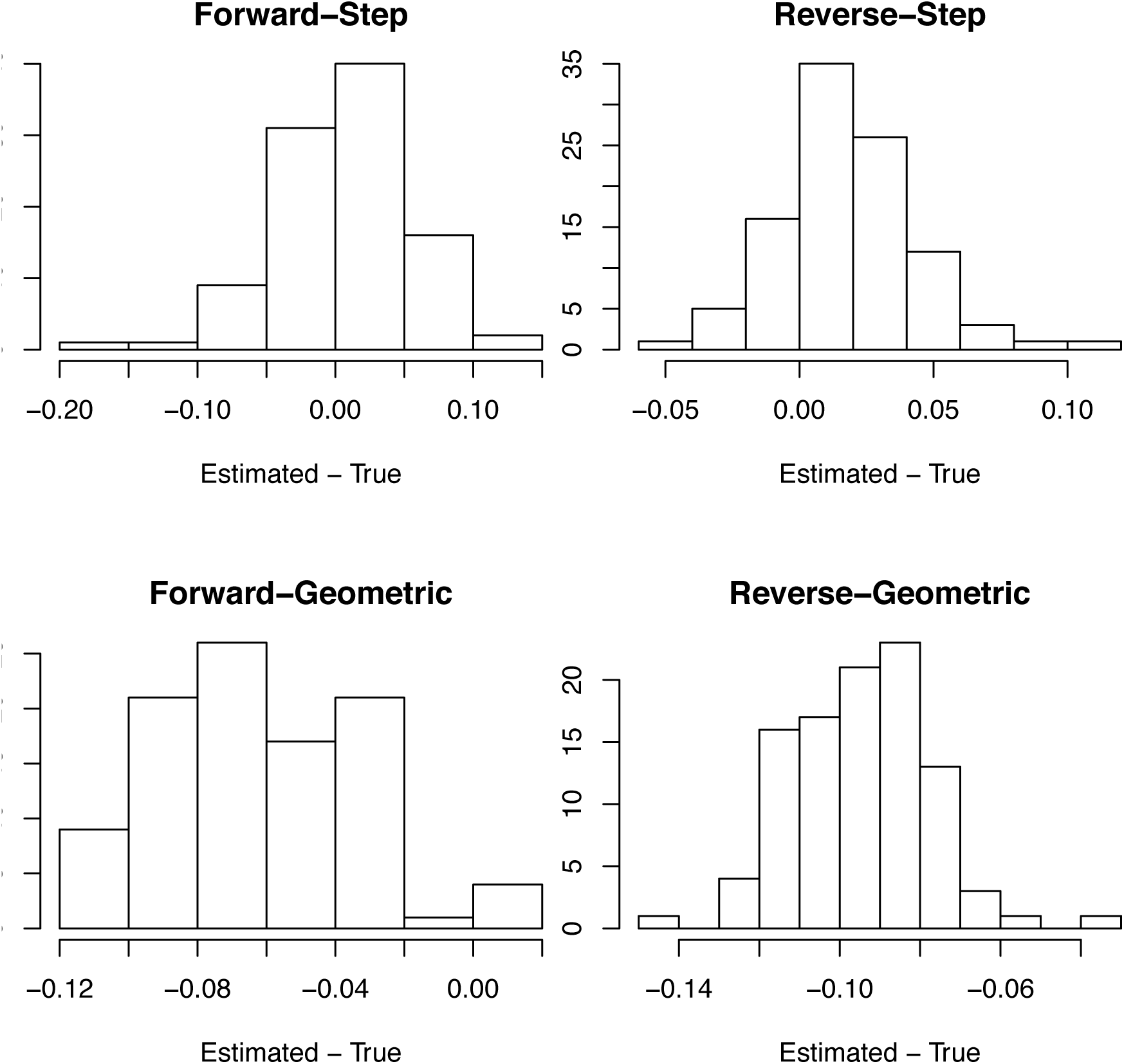
For case data, bias in the bootstrap estimates of *r*_0_ as the difference between the estimated and true values of *r*_0_ in the simulations. See figure S11.

**Fig. S14.**
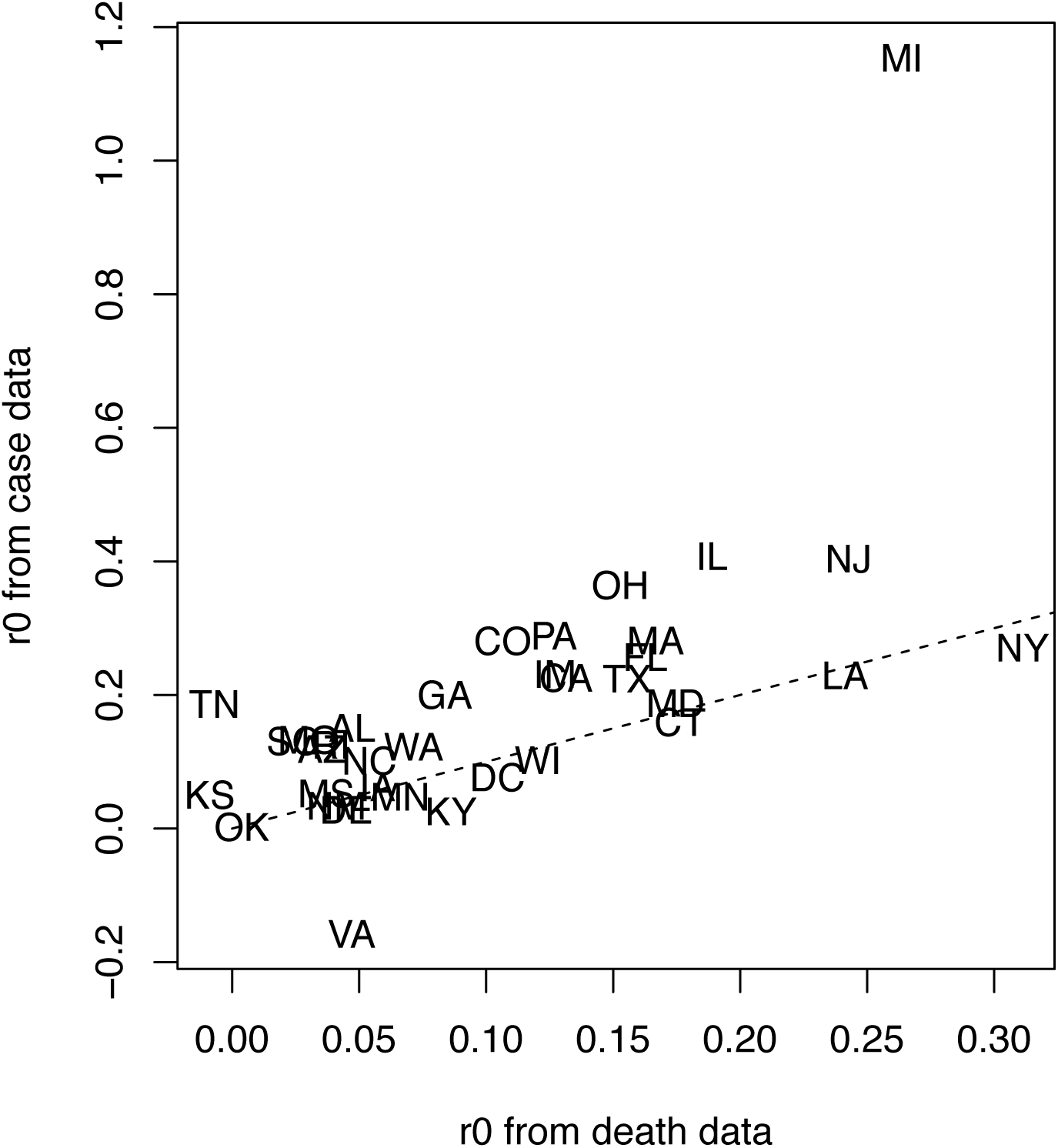
Comparison of *r*_0_ estimates from case data vs. death data.

**Fig. S15.**
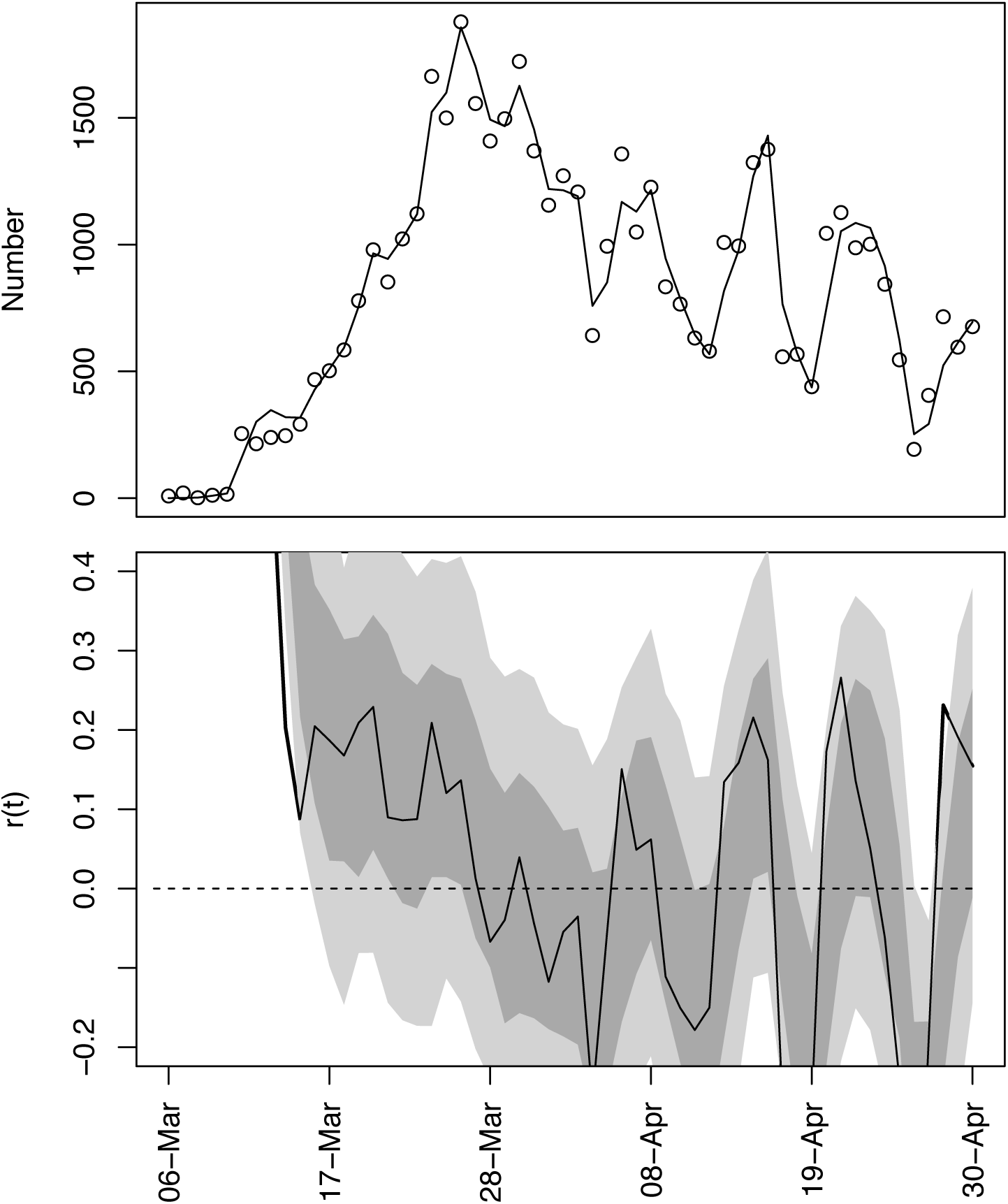
Case data and estimated *r*(*t*) for the State of Michigan. In (B), the poor fit is presumably caused by the high variance estimated for in ω*_r_*(*t*) (Eq. S1b) due to the weekly cycles in reported cases.

